# Improving Practical Diagnostic Measure for Enteric Parasites: Development of Fluorescent Antibody Microscopy (FAM) for Giardiasis and Cryptosporidiosis

**DOI:** 10.1101/2025.08.01.25332590

**Authors:** Yusuke Oshiro, Akira Kawashima, Megumi Akashi, Yasuaki Yanagawa, Kanako Shimodaira, Haruka Matsuzawa, Takashi Nemoto, Rieko Shimogawara, Masami Kurokawa, Naokatsu Ando, Haruka Uemura, Takahiro Aoki, Kei Yamamoto, Junichi Akiyama, Daisuke Mizushima, Toshio Kitazawa, Hiroyuki Gatanaga, Kenji Yagita, Koji Watanabe

**Author notes:** Corresponding authors: Koji Watanabe (corresponding author), Department of Parasitology, Division of Host Defense Mechanism, Tokai University, School of Medicine, 143 Shimokasuya, Isehara, Kanagawa 259-1193, Japan. These authors contributed equally to the present study (Yusuke Oshiro, Akira Kawashima).

## Abstract

**Background:** Giardiasis and cryptosporidiosis are often misdiagnosed by stool ova and parasite test (classical O&P). Multiplex PCR and rapid antigen immunochromatography (rapid-IC) could offer high accuracy; however, its cost and restricted coverage, especially for parasites, limit their use. This study evaluated the efficacy of newly developed “fluorescent antibody microscopy (FAM)” for these protozoa.

**Methodology/Principal Findinges:** DyLight 488-conjugated antibodies against *Giardia* cysts and *Cryptosporidium* oocysts were used for FAM examination. Diagnostic accuracy was assessed by reference to multiplex PCR results and compared with that of rapid-IC. FAM identified *Giardia* and/or *Cryptosporidium* in 35 of 694 stool samples tested. Also, FAM-negative stools examined in a randomly chosen month (49 samples) were selected as negative control. For *Giardia*, all FAM results were identical to those of rapid-IC, including three false negatives [100% positive predictive value (PPV), and 95.2% negative predictive value (NPV)]. For *Cryptosporidium*, FAM showed 100% PPV, and 98.6% NPV, which were comparable with those of rapid-IC. Conventional PCR for *Giardia* sequences produced negative results for three samples considered to yield FAM false negative, raising another possibility of multiplex PCR false positives.

**Conclusions/Significance:** Concurrent use of FAM with classical O&P, which strengthens diagnostic accuracy for Giardiasis/Cryptosporidiosis, is an option for cost-effective, practical diagnosis of enteric parasites.

**Author summary:** Giardiasis and Cryptosporidiosis are often misdiagnosed through traditional stool tests. Although multiplex-PCR and rapid antigen tests (rapid-IC) offer high accuracy, their cost and limited parasite coverage restrict their use. To overcome these challenges, we developed a new diagnostic method named “Fluorescent Antibody Microscopy (FAM).” This study evaluated the diagnostic accuracy of FAM by using multiplex-PCR as a reference standard and compared it with rapid-IC. For Giardia, FAM’s results were identical to rapid-IC, including three false negatives (100% PPV and 95.2% NPV). Similarly, for Cryptosporidium, FAM showed high diagnostic accuracy (100% PPV and 98.6% NPV), comparable to rapid-IC. Furthermore, in some cases, the concomitant use of bright-field microscopic examination occasionally identified enteric parasites, which were not covered by multiplex-PCR. Our results highlight that morphological parasitical identification with FAM may provide a cost-effective diagnostic strategy for enteric parasites, offering high accuracy for Giardia and Cryptosporidium.

## Introduction

Diarrheal diseases affect approximately 4 billion people worldwide annually, with waterborne protozoan infections being a major cause [1–4]. Protozoan infections, such as giardiasis and cryptosporidiosis, are primarily transmitted via the fecal–oral route in situations of poor sanitation, inadequate water supply, and environmental contamination [5–7]. Although these infections are mainly prevalent in developing countries, they can occur in developed countries as sporadic cases or because of an outbreak [4, 6, 8].

In Japan, giardiasis and cryptosporidiosis are classified as “Category V infectious diseases” under the Act on the Prevention of Infectious Diseases and Medical Care for Patients with Infectious Diseases mandating nationwide reporting of all confirmed cases [9, 10]. Surveillance data indicate international travelers and HIV-infected individuals to be at high risk for these protozoan infections [6, 11]. In addition, notable outbreaks of *Cryptosporidium* infection have been reported since its classification as a notifiable disease [10]. Moreover, diarrheal symptoms caused by these protozoa are indistinguishable from viral or bacterial gastroenteritis; therefore, these protozoa are frequently overlooked in common clinical settings [4, 5]. Their diagnosis should be improved to enhance patient care and to enable prompt epidemiological actions to protect public health.

The diagnosis of *Giardia* and *Cryptosporidium* currently relies on three main methods: morphological identification by classical ova and parasite bright-field microscopy (classical O&P), antigen detection, and polymerase chain reaction (PCR) [3–5, 7]. This is because intestinal protozoa cannot be cultivated in the microbiology laboratories of medical facilities [3, 4]. Classical O&P is highly affected by the skill of the examiner and is rarely used in most developed countries because of low diagnostic accuracy [4, 5]. Conversely, multiplex PCR has been increasingly used for diagnosis and research purposes because it offers high sensitivity and specificity [11, 13–16]. It can also detect numerous enteric pathogens, including bacteria, viruses, and protozoa in the same assay. However, multiplex PCR is expensive (∼$100 per test); therefore, its use is highly limited to specialized laboratories. Another common approach is rapid antigen detection by immunochromatography (rapid-IC), which provides rapid and user-friendly results without requiring specialized expertise [8, 17]. However, its relatively high cost (∼$30 per test for the detection of only a few pathogens) remains a concern for routine testing of enteric infectious diseases. Moreover, multiplex PCR and rapid-IC only target pre-defined pathogens, which is a crucial limitation in the diagnosis of intestinal parasites. Diagnosis of many protozoa as well as most helminths depend on classical O&P in clinical settings, although nucleic acid identification of each parasite can be applied for research purposes. Furthermore, identification by classical O&P is not difficult for most intestinal parasites except for small protozoa (< 10 μm). Thus, classical O&P has the advantage of being usable to examine various intestinal parasites within a routine protocol without incurring additional cost. Improving the diagnostic accuracy of O&P, for example by enabling easier identification of frequently overlooked common but small-sized protozoa, such as *Giardia* and *Cryptosporidium*, is a possible solution for enhancing the clinical diagnosis of intestinal parasites.

Here, we examined fluorescent antibody microscopy (FAM) in combination with classical O&P for the diagnosis of intestinal parasites. FAM assays enable morphological identification of *Giardia* and *Cryptosporidium* by fluorescence microscopy after the treatment of samples with DyLightⓇ488-conjugated antibodies specific for these protozoa. Diagnostic values for the targeted protozoa were assessed relative to multiplex PCR results and were compared with rapid IC results.

## Methods

### Sample collections and study approval

Stool samples submitted for microbiological examination from patients with suspected infectious gastroenteritis were collected between January 2022 and April 2025 (Fig. 1). Samples of sufficient volume for FAM, rapid-IC, and multiplex PCR were selected. These diagnostic tests, as well as DNA extraction using the QIAamp PowerFecal Pro DNA Kit© (Qiagen, Hilden, Germany), were performed immediately after sampling. Extracted DNA was stored at −80°C. Microbiology results were the only patient clinical information used. Informed consent was not obtained because all submitted specimens were anonymized; however, an opt-out was employed in accordance with ethical guidelines. This study was approved by the Institutional Review Board of the National Center for Global Health and Medicine (NCGM-S-004414).

**Figure 1.**
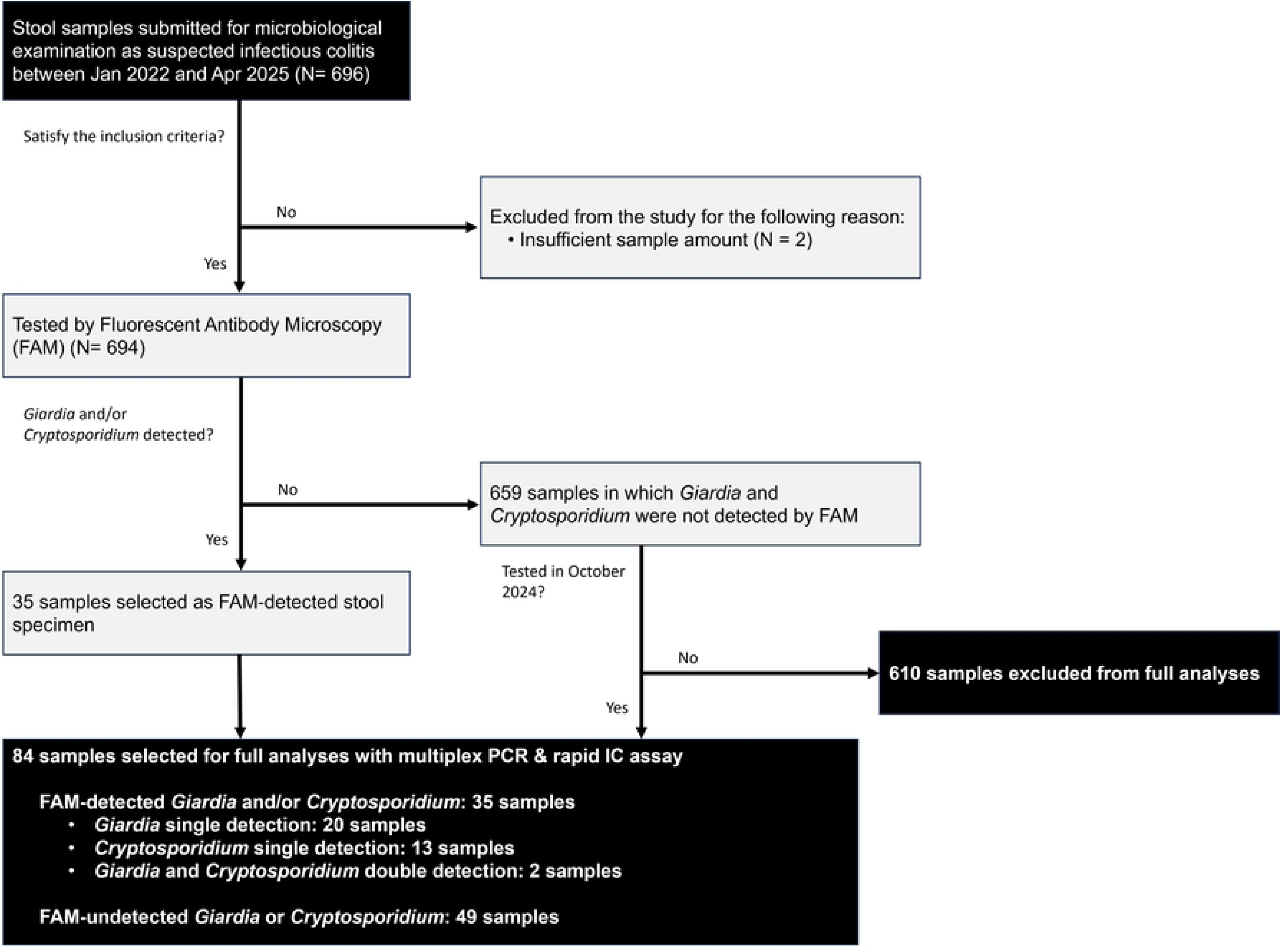
Sample collection. This flowchart illustrates the selection process of stool samples analyzed in the present study. Between January 2022 and April 2025, 696 stool samples were submitted for microbiological examination. Among them, 694 samples were included for analysis, with two excluded. Thirty-five samples tested positive for *Giardia* and/or *Cryptosporidium* by fluorescent antibody microscopy (FAM-positive samples), and an additional 49 FAM-negative samples submitted in October 2024 were selected as controls. These 84 samples underwent full analysis, including multiplex PCR and rapid immunochromatographic (IC) testing.

### Fluorescent Antibody Microscopy (FAM) for Giardia and Cryptosporidium

Fluorescent staining prior to morphological diagnosis was performed using ARK Checker C/G - DyLight 488 (ARK Resource, Japan). This reagent contains DyLight 488-conjugated monoclonal IgG antibodies targeting *G. duodenalis* cysts and *Cryptosporidium* oocysts, and was originally designed by our group to test the quality of the water supply system in Japan [18]. Specimens were prepared using the formalin-ether sedimentation method. The sedimented fraction (10 μL) was mixed with 10 μL of ARK Checker C/G - DyLight 488 and mounted on a glass slide under a coverslip.

Fluorescence images were obtained through a 510–550-nm wavelength filter after B-excitation (490 nm) (Fig. 2). Four clinical laboratory technologists, who had no experience of fluorescence microscopy, performed FAM diagnosis. They underwent a 1-hour training session using positive control samples of *G. duodenalis* and *Cryptosporidium* spp. prior to the study. Diagnosis was made by a single examiner during the study period.

**Figure 2.**
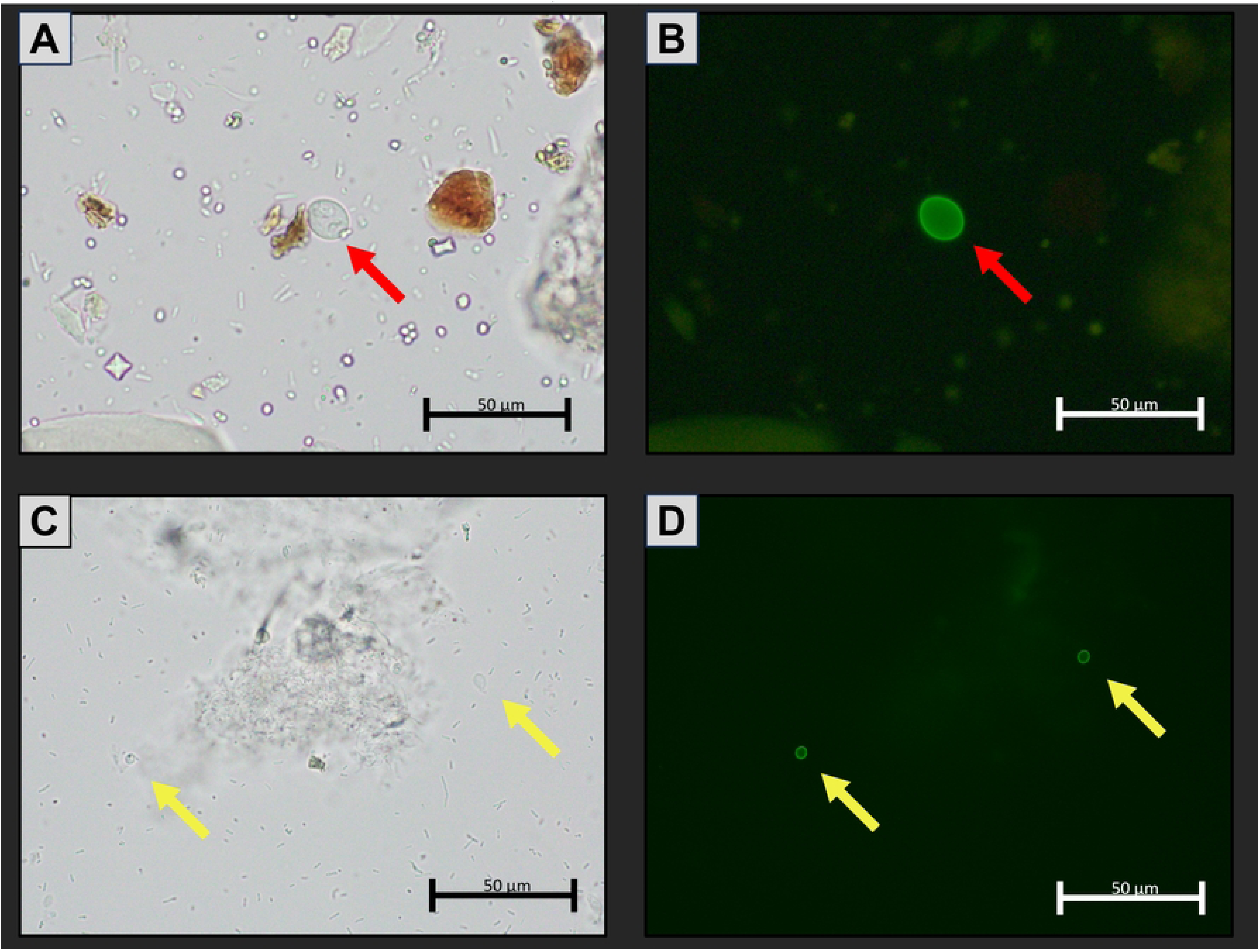
Bright-field microscopy and fluorescent antibody microscopy (FAM) images of *Giardia* and *Cryptosporidium*. Bright-field microscopy image (A), and FAM image (B) of *Giardia* cysts, indicated by red arrows. Bright-field microscopy image (C), and FAM image (D) of *Cryptosporidium* oocysts, indicated by yellow arrows. FAM images were obtained through an absorption filter (510–550 nm) after excitation with a blue laser (490 nm).

### Rapid Immunochromatographic (rapid-IC) Assays, and Multiplex PCR

Rapid-IC was performed using a GIARDIA/CRYPTOSPORIDIUM QUIK CHEK test (TECH LAB, USA), and multiplex PCR was performed using a Multiplex PCR FilmArray Gastrointestinal Panel (BioMérieux, France) according to the manufacturer’s instructions.

### Sequence analysis of discordant cases

For cases with discrepancies among FAM, rapid IC assay, and multiplex PCR results, we performed PCR followed by Sanger sequencing using DNA extracted from stool samples. For *Giardia duodenalis*, nested PCR targeting the glutamate dehydrogenase (*gdh*) gene was performed. Outer primers Ghd1 (5′-TTCCGTRTYCAGTACAACTC-3′) and Gdh2 (5′-ACCTCGTTCTGRGTGGCGCA-3′), and nested primers Gdh3 (5′-ATGACYGAGCTYCAGAGGCACGT-3′) and Gdh4 (5′-GTGGCGCARGGCATGATGCA-3′) were used [19]. For *Cryptosporidium* spp., conventional PCR targeting the 18S rRNA gene was conducted using forward (5′-AAGCTCGTAGTTGGATTTCTG-3′) and reverse (5′-TAAGGTGCTGAAGGAGTAAGG-3′) primers [20]. The cycling conditions were as follows: an initial denaturation at 94°C for 3 minutes, followed by 40 cycles of denaturation at 94°C for 30 seconds, annealing at 60°C for 30 seconds, and extension at 72°C for 30 seconds. The nested *Giardia* PCR used an annealing temperature of 67°C and 25 cycles. Sequence analysis of each amplicon was performed by Sanger sequencing using appropriate forward and reverse primers (Eurofins Genomics, Tokyo, Japan).

### Statistical analysis

Statistical analyses were performed using GraphPad Prism (v10.2.3). Sensitivity, specificity, positive predictive value (PPV), and negative predictive value (NPV) with 95% confidence intervals (Cis) were calculated for FAM and rapid IC using multiplex PCR as a reference. Fisher’s exact test was used to compare FAM and rapid IC results, with statistical significance set at p < 0.05.

## Results

### Sample collection

During the 3 years of the study period, 696 stool samples collected from patients with suspected infectious colitis were examined (Fig. 1). Of these, two samples were excluded from the analysis because of insufficient sample volume. Therefore, FAM was performed on 694 samples. We identified 35 stool samples positive for *Giardia* and/or *Cryptosporidium*, which we classified as FAM-positive stool samples. These protozoa were not identified in the other 659 samples. As an unbiased collection of FAM-negative stool samples for use as control samples, we chose all the FAM-negative samples examined in 1 month, October 2024. Overall, we therefore selected 84 samples, 35 FAM-positive and 49 FAM-negative samples, for evaluating the diagnostic accuracy of three methods (Supplementary data 1).

### Diagnostic value of FAM

Diagnostic accuracy of FAM for *Giardia* and *Cryptosporidium* was evaluated by comparison with multiplex PCR (FilmArray Gastrointestinal Panel©) as a reference. Thereafter, it was compared with rapid-IC tests (GIARDIA/CRYPTOSPORIDIUM QUIK CHEK^©^ tests).

### Performance of FAM for Giardia

The sensitivity, specificity, positive predictive value (PPV), and negative predictive value (NPV) of FAM are shown in Table 1A. For all FAM-positive giardiasis cases, *Giardia lamblia* (syn. *G. duodenalis*) was detected by multiplex PCR (100% PPV). *G. lamblia* was also detected by multiplex PCR in three FAM-negative samples, suggesting false negative results of FAM (95.2% NPV). The diagnostic performance of rapid-IC was similarly assessed (Table 1B). PPV, NPV, sensitivity, and specificity of rapid-IC were all the same as those of FAM. More surprisingly, the same three samples showed false negative results for FAM and rapid-IC. These results indicate that diagnostic accuracy of FAM is comparable to that of rapid-IC. They also indicate that the false negative results for FAM and rapid-IC probably occurred because the pathogen burden was beneath the level of detection by these tests.

**TABLE 1.** Sensitivity and specificity of each method for the detection of *Giardia* with reference to multiplex PCR.

**A. Performance of fluorescent antibody microscopy examination**
| Giardiasis diagnosis relative to multiplex PCR<sup>a</sup> | Performance relative to PCR (no. of samples) |  |  | PPV (% [95% CI<sup>d</sup>]) | NPV (% [95% CI]) |
| --- | --- | --- | --- | --- | --- |
|  | Positive | Negative | Total |  |  |
| Fluorescent antibody microscopy examination |  |  |  |  |  |
| Positive | 22 | 0 | 22 | 100 (85.1–100) |  |
| Negative | 3 | 59 | 62 |  | 95.2 (86.7–98.7) |
| Total | 25 | 59 |  |  |  |
| Sensitivity (% [95% CI<sup>d</sup>]) | 88.0 (70.0–95.8) |  |  |  |  |
| Specificity (% [95% CI<sup>d</sup>]) |  | 100 (93.9–100) |  |  |  |

**B. Performance of rapid IC test**
| Giardiasis diagnosis relative to multiplex PCR<sup>a</sup> | Performance relative to PCR (no. of samples) |  |  | PPV (% [95% CI<sup>d</sup>]) | NPV (% [95% CI]) |
| --- | --- | --- | --- | --- | --- |
|  | Positive | Negative | Total |  |  |
| Rapid IC test <sup>b</sup> |  |  |  |  |  |
| Positive | 22 | 0 | 22 | 100 (85.1–100) |  |
| Negative | 3 | 59 | 62 |  | 95.2 (86.7–98.7) |
| Total | 25 | 59 |  |  |  |
| Sensitivity (% [95% CI<sup>d</sup>]) | 88.0 (70.0–95.8) |  |  |  |  |
| Specificity (% [95% CI<sup>d</sup>]) |  | 100 (93.9–100) |  |  |  |

<sup>a</sup> Performed using a FilmArray Intestinal Panel <sup>b</sup> CI, confidence interval. <sup>c</sup> Immunochromatography by GIARDIA/CRYPTOSPORIDIUM QUIK CHEK test.

### Performance of FAM for Cryptosporidium

The performance of FAM for detecting *Cryptosporidium* is shown in Table 2A. *Cryptosporidium* spp. was detected by multiplex PCR in all FAM-positive samples, and in one sample among 64 FAM-negative samples (100% PPV, 98.6% NPV) (Table 2). In addition, rapid-IC produced no false-positive results for *Cryptosporidium*. However, rapid-IC produced false negative results in two cases (100% PPV, 97.1% NPV), one of which was the sample that gave a false negative result by FAM. These results indicate the diagnostic accuracy of FAM for *Cryptosporidium* to be highly comparable to that of rapid-IC.

**TABLE 2.**
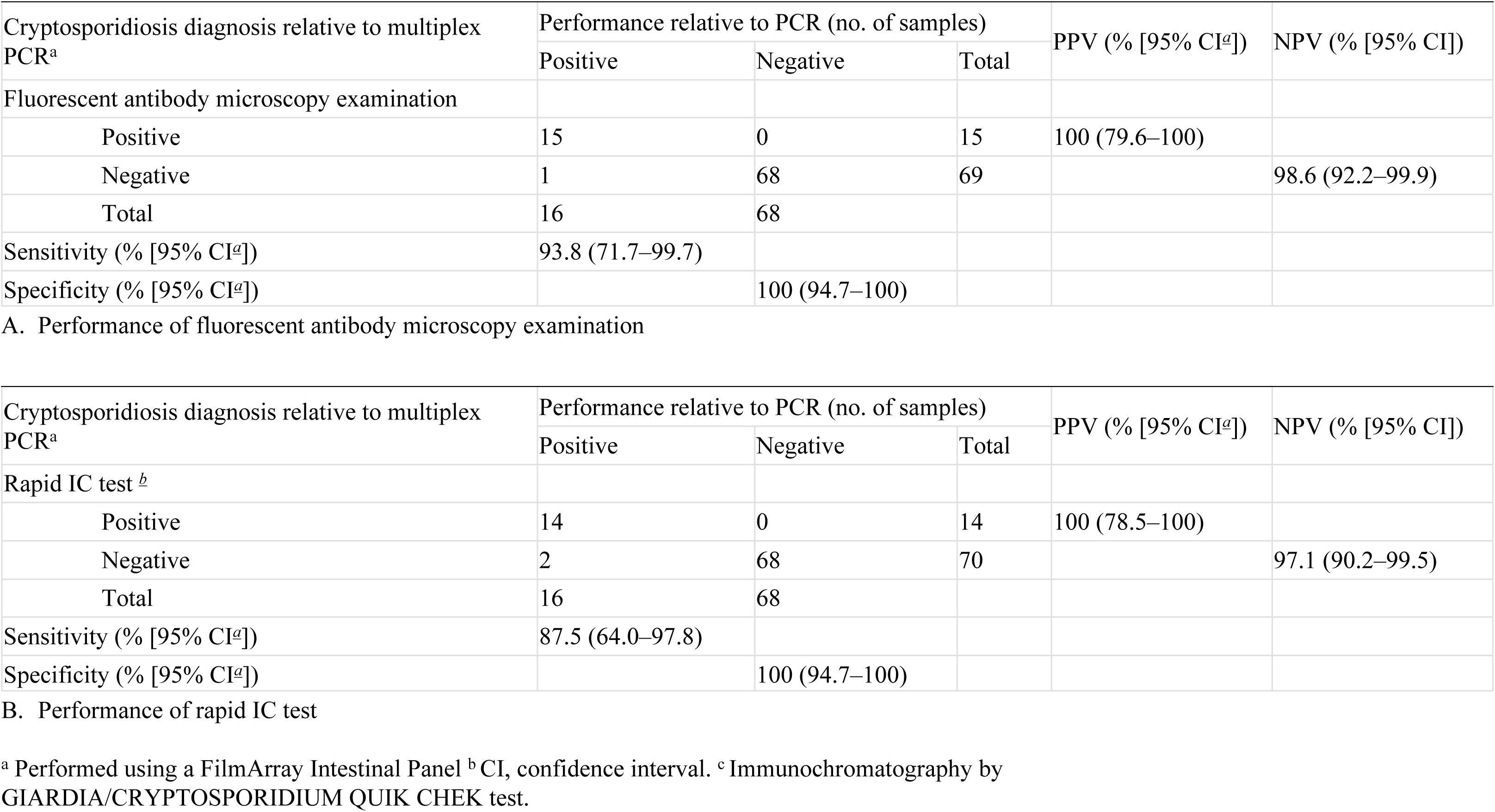
Sensitivity and specificity of each method for the detection of *Cryptosporidium* with reference to multiplex PCR.

### Evaluation of Discordant Cases Using Nested and Conventional PCR

As described above, discordant (false positive/negative) results produced by the different testing methods were seen in five cases, three discordant for *Giardia* and two discordant for *Cryptosporidium* (Table 3). As already mentioned, the cases discordant for *Giardia* yielded the same results by three diagnostic measures; they were FAM/rapid-IC-negative but multiplex PCR-positive. To confirm the presence of these pathogens in these samples, we performed conventional or nested PCR with sequencing analysis.

**TABLE 3.**
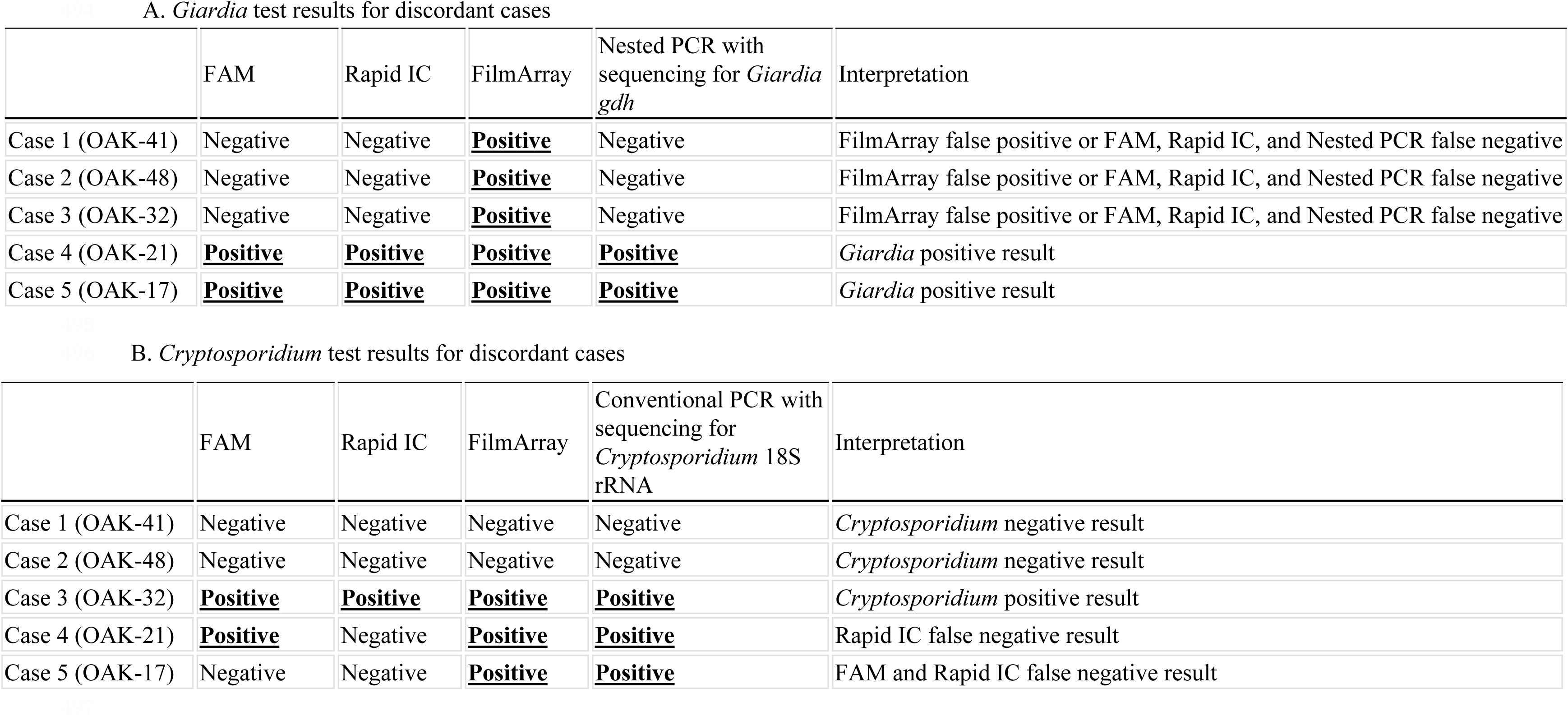
Analysis of discordant cases among different diagnostic methods.

Notably, in all samples discordant for *Giardia*, *G. duodenalis* could not be identified by nested PCR (Table 3A, Case 1-3), whereas *G. duodenalis* was clearly identified in the *Giardia*-positive sample (Table 3A, Case 4&5). These results raised two possibilities for the discordant *Giardia* results; one is an extremely low pathogen burden of *G. duodenalis*, resulting in a positive result only by multiplex PCR, and the other is a false positive multiplex PCR result.

For cases with discordant *Cryptosporidium* results, we performed conventional PCR, which produced the same results as multiplex PCR (Table 3B). These findings confirm that multiplex PCR reliably identified *Cryptosporidium* spp. in the present analysis.

## Discussion

Improved treatment of intestinal parasitic diseases requires a low-cost, rapid, and technically simple diagnostic measure covering various parasites. In addition, the method should be developed as highly sensitive to small-sized common protozoa, such as *Giardia* and *Cryptosporidium*, which are frequently overlooked by classical O&P. Multiplex PCR and antigen testing show high sensitivity to these protozoa [8, 14, 17, 21]; however, these tests are generally expensive. Moreover, both tests can detect only pre-defined pathogens; therefore, other “untargeted” parasites need to be diagnosed separately. Currently, some intestinal protozoa and most helminths are not recognized by multiplex PCR or rapid IC, and need to be identified morphologically by classical O&P in clinical settings. In the present study, we examined FAM using fluorescence-conjugated specific antibodies against *G. duodenalis* and *Cryptosporidium* spp. for increased sensitivity to these protozoa, as an auxiliary diagnosis method to classical O&P.

First, FAM exhibited high specificity for *Giardia*, comparable to that of rapid IC. Additionally, the sensitivity of FAM was the same as that of rapid-IC. However, FAM and rapid-IC produced false negative results for *Giardia* in the same samples relative to the reference results of multiplex PCR. *G. duodenalis* was not identified by nested PCR targeting *gdh*. These results indicate that the pathogen burden was too low to be detected by these methods but not by multiplex PCR, or that multiplex PCR produced a false positive result. The manufacturer of the Multiplex PCR FilmArray Gastrointestinal Panel notes cross-reactivity between *Giardia* and other gut microbiota, such as *Bifidobacterium* spp. and *Ruminococcus* spp. [22]. In addition, multiplex PCR frequently reports multiple pathogens in one stool specimen, some of which do not match the results of other diagnostic methods, such as bacterial culture [23]. In fact, in the present study, half of FAM-positive “*Giardia* and/or *Cryptosporidium* containing” samples (18 out of 35) showed multiple pathogens by multiplex PCR (*G. lamblia*/*Cryptosporidium* spp. plus one or more other microorganisms were positive simultaneously. Supplementary data 1). Such multiplex PCR results often make it difficult for physicians to determine the causative agent of intestinal disease. Considered together, FAM has good diagnostic value for giardiasis and is comparable to that of rapid IC.

We emphasize that FAM produced only one false negative case for *Cryptosporidium*, whereas rapid-IC produced an additional false negative case. Additionally, FAM did not produce any false positives. In general, identification of *Cryptosporidium* oocysts is extremely difficult because of their relatively small size compared with other protozoa. We predict that FAM can decrease the number of overlooked or misdiagnosed cases of cryptosporidiosis that frequently occur by classical O&P examination.

The effectiveness of fluorescence-based diagnostic tools has already been applied to other organisms [24]. For example, auramine staining has been used for tuberculosis screening (excitation: 460 nm, emission: 550 nm) [25, 26]. Accordingly, microbiology laboratories in most regional hospitals in Japan are equipped with fluorescence microscopy capability. The running cost of FAM is extremely low (up to $2 per test) compared with that of multiplex PCR or rapid IC assays. In addition, FAM can be simultaneously performed with classical O&P. In fact, tested stool samples contained some enteric parasites (e.g. *Sarcocystis* spp. and *Taenia saginata*), which were uncovered by multiplex PCR (OAK-40 & −48 in Supplementary data 1). They were morphologically diagnosed by classical O&P. Furthermore, *Cyclospora cayetanensis* can be identified by fluorescent microscopy through its autofluorescence by ultraviolet excitation (supplementary data 2). Based on these findings, we suggest a FAM-containing diagnostic algorithm for enteric parasites (Fig. 3). First, FAM with classical O&P is employed to screen for enteric parasites when infectious gastroenteritis is suspected. Its implementation could enhance diagnostic accuracy while minimizing the risk of overlooking common protozoa and maintaining cost-effectiveness. Rapid-IC and/or multiplex PCR should be performed in limited cases to narrow down differential diagnoses (e.g. ruling out of infectious etiology before diagnosing non-infectious inflammatory bowel diseases).

**Figure 3.**
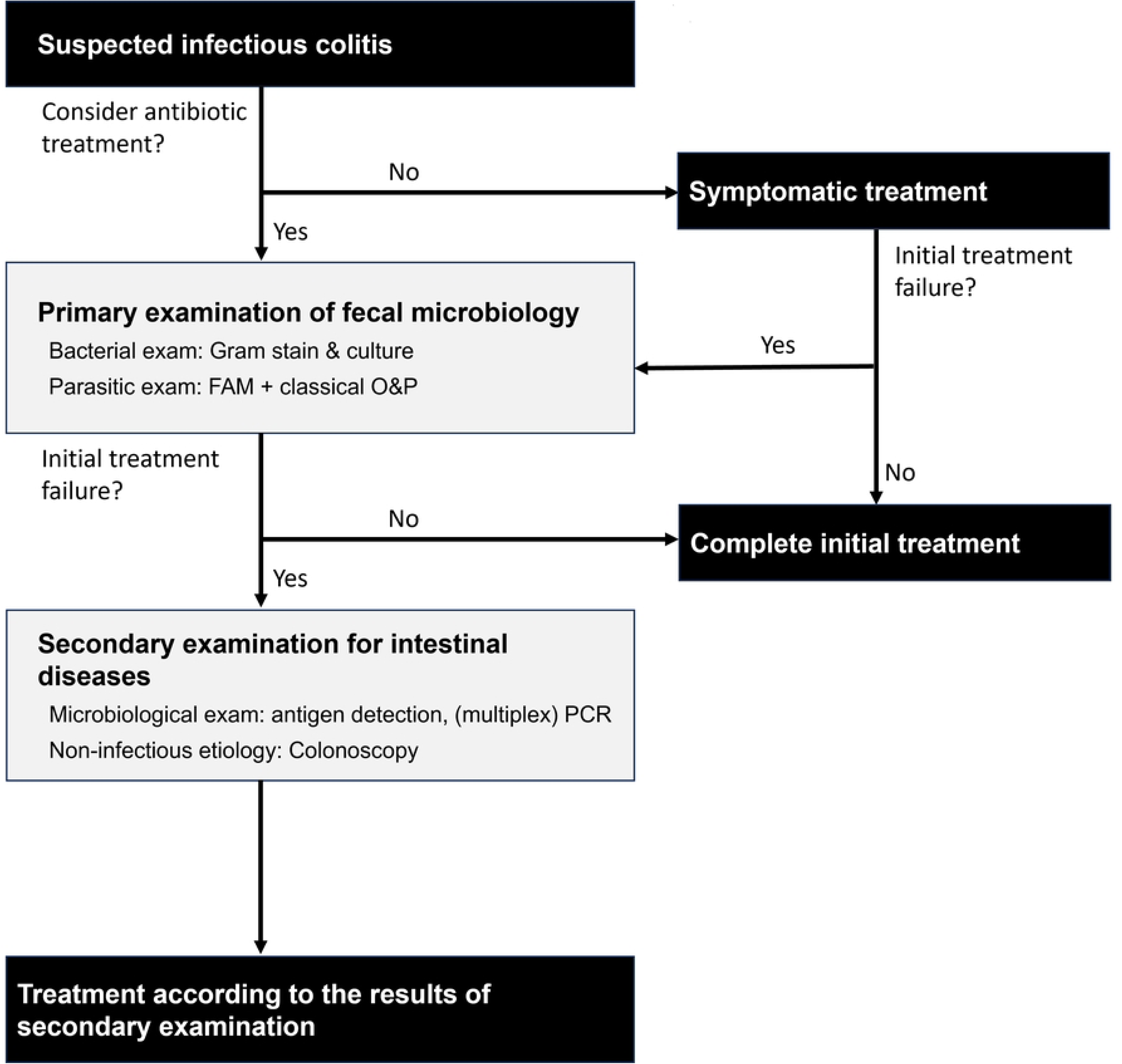
Proposed diagnostic flowchart for infectious enteritis including protozoa detection by FAM. This diagram illustrates a suggested diagnostic algorithm for infectious colitis. Initial screening tests include Gram staining and culture for bacterial pathogens, as well as fluorescent antibody microscopy (FAM) combined with classical ova and parasite bright-field microscopy (classical O&P) for intestinal protozoa and helminths. If no causative agents are identified despite persistence of clinical symptoms, rapid immunochromatographic (IC) assays or multiplex PCR testing are recommended as secondary tests. This approach aims to optimize diagnostic accuracy while maintaining cost-effectiveness and minimizing overlooked protozoan infections, such as giardiasis and cryptosporidiosis.

This study has some limitations. First, it was conducted as a single-center study at a general hospital in Tokyo, which serves as a national reference center for travel and HIV-related infections. The number of intestinal parasite cases seen is relatively high compared with other facilities in Japan; however, positive cases of *Giardia* and/or *Cryptosporidium* are still limited. In addition, the small number of examiners from the center involved in the study were more experienced in classical O&P than general practitioners. Therefore, diagnostic accuracy could be overestimated. Considered together, a larger scale study conducted at multiple facilities in resource limited areas is warranted for more reliable assessment of the diagnostic utility of FAM.

In conclusion, we demonstrated that FAM has comparable diagnostic accuracy for *Giardia* and *Cryptosporidium* compared with rapid-IC while having the advantages of cost-effectiveness, examiner independence, and practical feasibility. Continuous efforts to improve the diagnostic flow for enteric parasites are needed for better clinical practice.

## Acknowledgments

We thank Jeremy Allen, PhD, from Edanz (https://jp.edanz.com/ac) for editing a draft of this manuscript.

All authors contributed significantly to the study. Y.O., K.Yagita, and K.W. were responsible for the study overall and for study design and conceptualisation. Y.O., A.K., and K.W. wrote the manuscript. Y.O., E.A., K.S., and T.K. performed the fluorescent antibody microscopy and rapid immunochromatographic assay. A.K., R.S., M.K., and K.Yagita performed the PCR analysis. A.K., Y.Y., N.A., T.A., and D.M. recruited participants and managed clinical data collection. H.U., K.Yamamoto, J.A., and H.G. supervised the study. Figures and tables were prepared by A.K. and Y.O. Funding was acquired by A.K. and K.W. All authors approved the final version of the manuscript.

## Ethical Approval Statement

This study was approved by the Institutional Review Board of the National Center for Global Health and Medicine (approval number: NCGM-S-004414). Informed consent was not obtained because all submitted specimens were anonymized; however, an opt-out approach was employed in accordance with institutional ethical guidelines.

## Data Availability

All data generated or analyzed during this study are included in this published article and its supplementary information files.

